# Assessment of common genetic variation in Alzheimer’s and Parkinson’s diseases reveals global distinction in population attributable risk

**DOI:** 10.1101/2024.09.23.24314240

**Authors:** Lietsel Jones, Catalina Cerquera-Cleves, Artur FS Schuh, Mary B Makarious, Hirotaka Iwaki, Mike A. Nalls, Alastair J Noyce, Global Parkinson’s Genetics Program (GP2), Cornelis Blauwendraat, Andrew Singleton, Ignacio Mata, Mark R. Cookson, Sara Bandres-Ciga

## Abstract

Emerging evidence suggests that the genetic architecture of Alzheimer’s (AD) and Parkinson’s diseases (PD) risk varies across ancestries. This study seeks to explore distinct and universal genetic targets across individuals of Latino, African/African Admixed, East Asian, and European populations by implementing Population Attributable Risk (PAR) comparisons on summary statistics from genome-wide association studies (GWAS). PAR was calculated for the most significant disease variants using summary statistics derived from select multi-ancestry GWAS meta-analyses, followed by fine-mapping analysis to validate genetic contribution of disease variants to European, African/African Admixed, East Asian, and Latino individuals. For both AD, *APOE4* PAR estimates were universally high across all ancestries, with *TSPAN14* and *PICALM* emerging as other common targets. Attributable risk varied across PD-related major risk loci including variation nearby *GBA1* and *LRRK2*. In contrast, *SNCA, MCCC1, VPS13C*, and *MAPT* loci demonstrated comparable attributable risk across ancestries. This cross-ancestry evaluation of PAR reinforces the genetic heterogeneity of AD and PD. In consideration of the complex etiology of these diseases, these findings may inform the strategic prioritization of therapeutic targets and improve global health outcomes.

## Introduction

Alzheimer’s (AD) and Parkinson’s (PD) have become increasingly prevalent worldwide with a growing elderly population contributing to high disease burden ^1^. Genome-wide association studies (GWAS) provide genetic evidence to better understand disease risk. Emerging global GWAS have uncovered key genomic regions associated with these conditions across different populations^2–10^.

Population Attributable Risk (PAR) is a statistical measure used in epidemiology to estimate the proportion of disease cases that would decrease if a risk factor was removed from a population ^11,12^. PAR quantifies impact of risk factors on disease occurrence, offering insights into disease pathophysiology and therapeutic strategies. Case-control GWAS meta-analyses report odds ratios (OR) as opposed to relative risk, and consequently, genetic PAR compares risk across genetic loci using OR and frequency of the most representative variant(s) in each region. PAR serves to prioritize genetic targets according to their impact on a specific population enabling healthcare strategies to be broadly applicable^12^.

Our aim was to estimate PAR for common genetic risk factors previously identified through multi-ancestry GWAS to prioritize distinct and universal therapeutic targets for precise applicability across genetically-defined ancestries.

## Methods

### Data sources

Reference datasets included summary statistics from published population-specific AD/PD GWAS meta-analyses across four genetically-defined ancestries with variable frequencies of many known risk factors across strata ^2–10^ (Table S1). We omitted proxy cases (not clinically defined cases) and focused on independent risk alleles. Information about data access and all scripts for analyses are publicly available on GitHub (DOI https://doi.org/10.5281/zenodo.13774455; https://github.com/GP2code/PAR-ADPD).

### Population Attributable Risk calculations

The most significant disease variants per locus were selected from recent multi-ancestry GWAS meta-analyses and we filtered out nominally significant variants (p < 0.05) to increase confidence in effect directionality and reduce confounding. OR and risk allele frequency (RAF) were calculated for single-nucleotide polymorphisms (SNPs) representing significant loci for each ancestry group. Variants with an OR > 1, (beta > 0) were classified as risk variants with RAF equal to the mean allele frequency (MAF) from summary statistics. For protective variants (OR < 1, beta < 0), the minor allele was flipped, and RAF was calculated as *RAF* = 1 − *MAF*.

We calculated PAR using the formula:

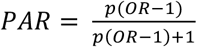

where p represents the RAF and OR is the calculated odds ratio for each risk allele (OR > 1, beta > 0). RAF values were plotted against their corresponding ORs for each genetic ancestry group.

Relevant risk loci were fine-mapped to validate genetic contribution of variants highlighted by the PAR method. Posterior probability greater than 80% suggested higher causality and disease burden. PAR estimates were ranked from highest to lowest values for cross-ancestry comparison.

## Results

A total of 55 AD and 90 PD variants from GWAS meta-analyses^2–10^ were evaluated as population-attributable risk factors for European, African/African Admixed, Latino, and East Asian predicted genetic ancestries (Table S2, Table S3). Table S4 highlights the variants with the highest PAR estimates for both AD and PD in each ancestry group.

### PAR comparisons reveal major AD risk locus APOE maintained high PAR across populations, while nominating other universally applicable genetic targets

*APOE* allelic variants (rs7412 and rs429358), consistently demonstrated the highest PAR and posterior probability across all groups compared to other variants (Figure 1)(Figure 3). Latino genetic ancestry individuals had the lowest ranked PAR estimates for *APOE* compared with other populations (Table S4).

**Figure 1.**
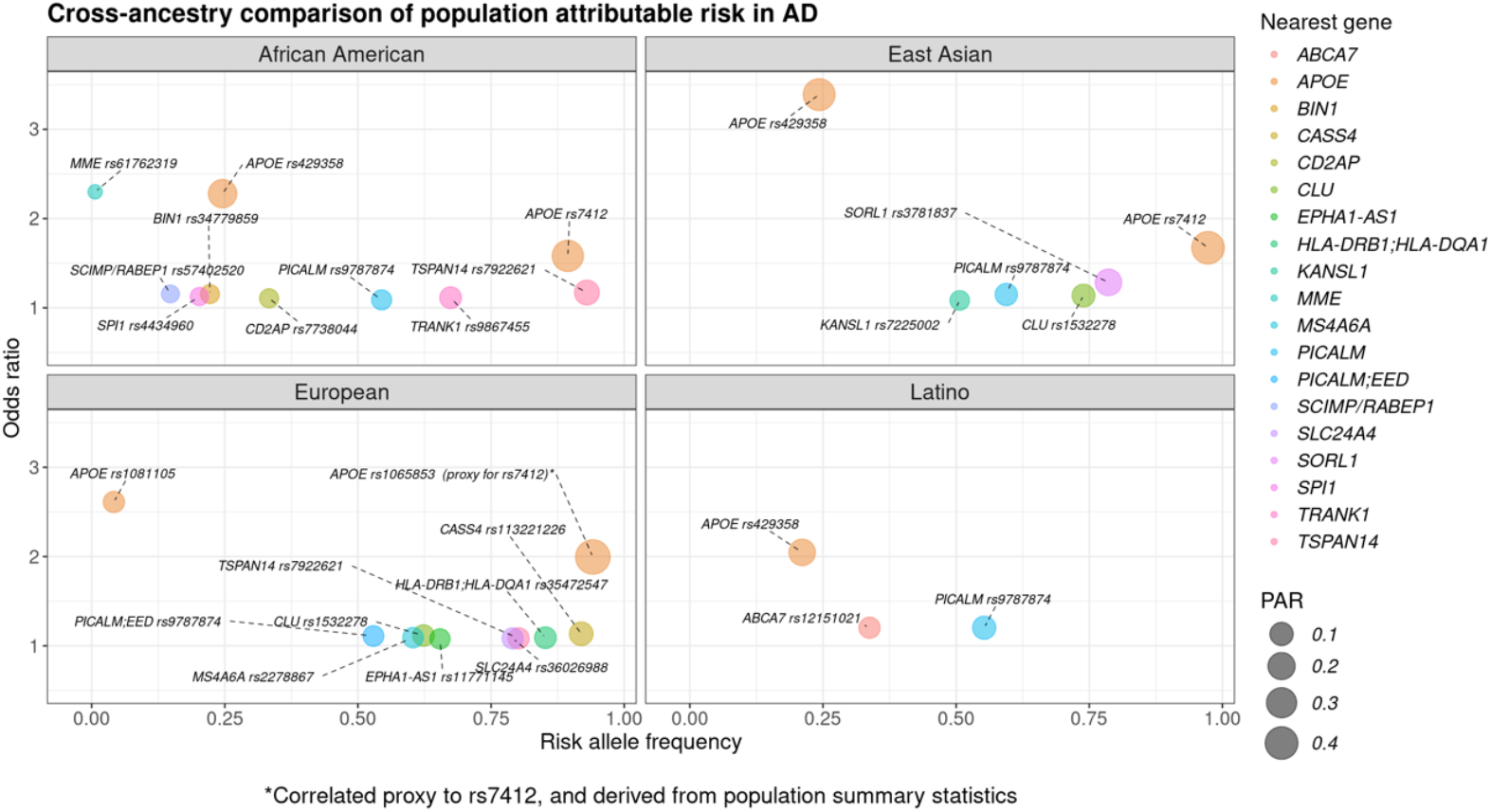
Comparison of population attributable risk (PAR) estimates for Alzheimer’s disease-related variants. Each panel highlights the variants with the highest PAR values in African American, East Asian, European, and Latino genetic ancestry strata, respectively. Color represents the nearest risk locus for assessed variants while the size corresponds to the PAR value. The risk allele frequency is reflected on the x-axis and the y-axis corresponds to the odds ratio, or effect size. As variant information was derived from population summary statistics, the *APOE* rs1065853 variant was used as a correlated proxy to APOE rs7412 in the European population.

Notably, AD-related locus *TSPAN14* represented by the rs7922621 variant exhibited one of the top signals in Europeans and African American genetic ancestry individuals. The *PICALM* rs9787874 variant had one of the highest PAR estimates across individuals from Latino, East Asian, African American, and European genetic backgrounds (Figure 1)(Table S4).

### Major PD-related loci LRRK2 and GBA1 exhibited variability in PAR estimates while other loci displayed cross-ancestry attributable risk

*GBA1* was identified as one of the top PAR signals for PD in the African/African Admixed genetic ancestry group with the highest posterior probability, primarily driven by the population-specific *GBA1* rs3115534 intronic variant (Figure 2)(Table S7). Coding variants in other ancestries demonstrated varying PAR estimates. PAR estimates for *GBA1* rs76763715 (p.N370S) were low in European and Latino genetic ancestries, as well as the estimates for the intronic *GBA1* rs146532106 variant tagging p.E326K in the East Asian strata of this study (Figure 3)(Table S3). Low PAR was observed in all groups for the *LRRK2* rs76904798 variant (p.G2019S), with the lowest occurring in Latino populations (Table S3).

**Figure 2.**
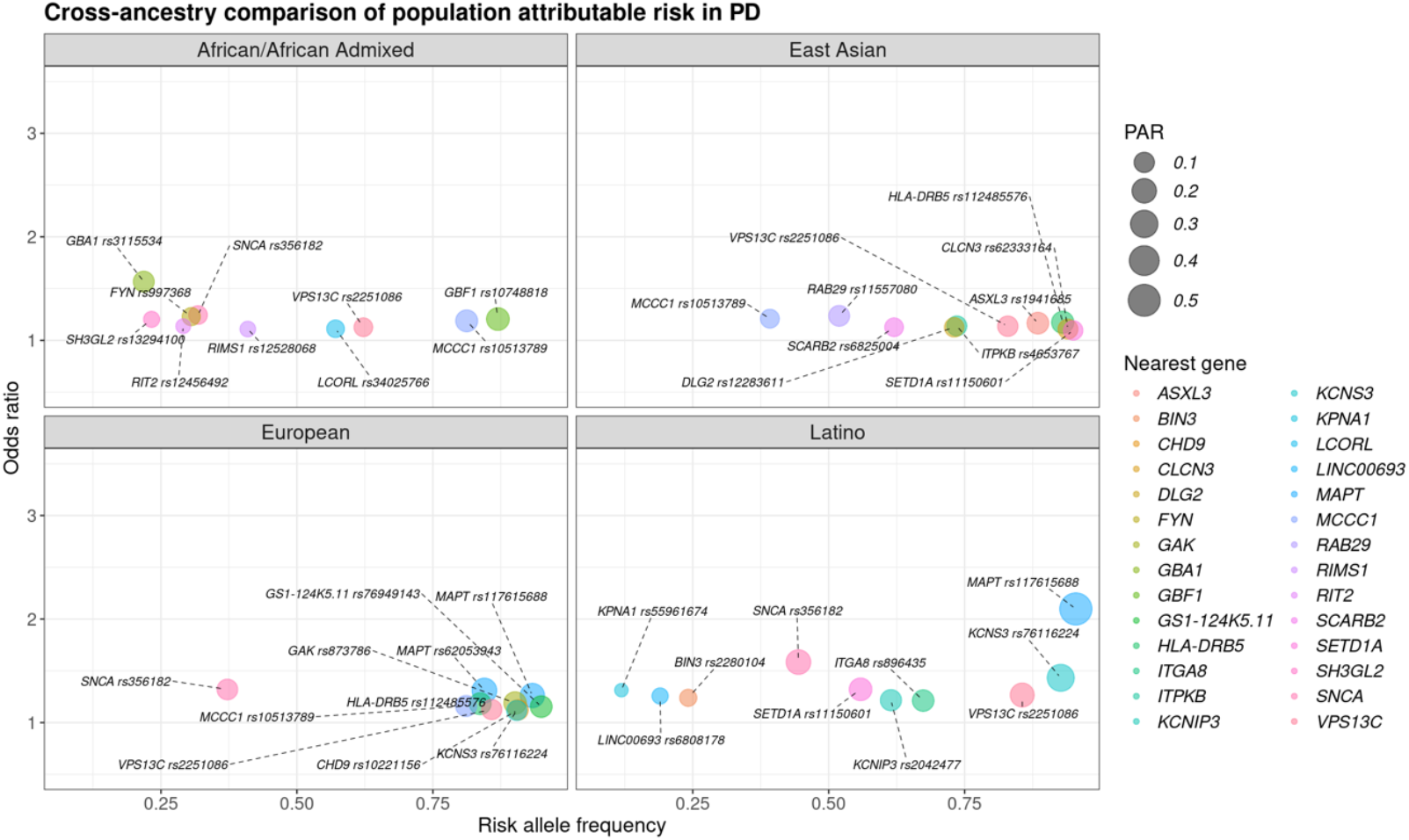
Comparison of population attributable risk (PAR) estimates for Parkinson’s disease-related variants. Each panel highlights the variants with the highest PAR values in African/African Admixed, East Asian, European, and Latino genetic ancestry samples, respectively. Color represents the nearest risk locus for assessed variants while the size corresponds to the PAR value. The risk allele frequency is reflected on the x-axis and the y-axis corresponds to the odds ratio, or effect size.

**Figure 3.**
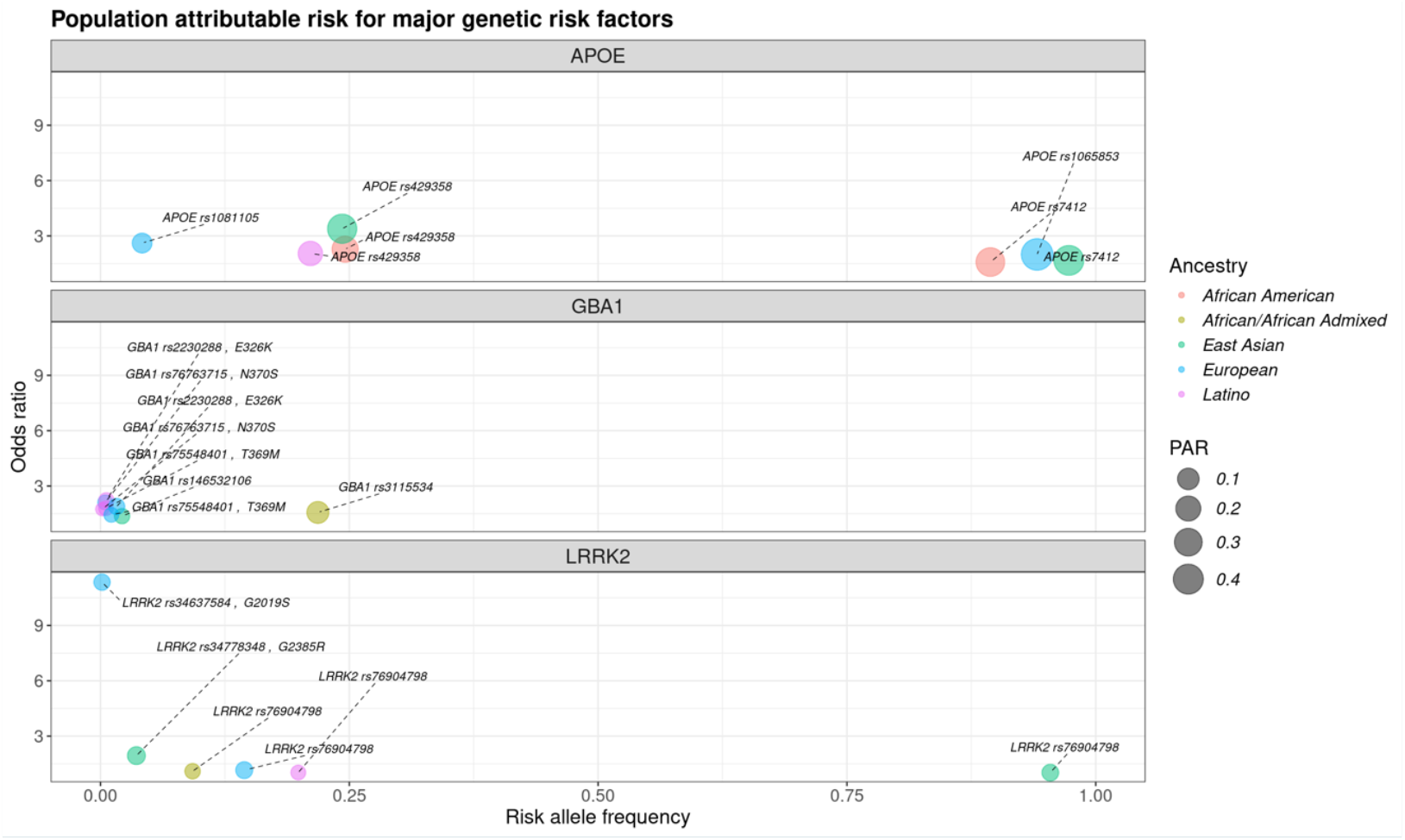
Population attributable risk (PAR) estimates for major known genetic risk factors. Each panel highlights PAR differences across populations for known genetic variants within *APOE, GBA1*, and *LRRK2*, respectively. Color represents the genetic ancestry strata while the size corresponds to the PAR value. The risk allele frequency is reflected on the x-axis and the y-axis corresponds to the odds ratio, or effect size.

The *MAPT* locus associated with PD had highest signal strength in Latino and European GWAS, while multi-signal *SNCA* had higher cross-ancestry estimates. A specific *SNCA* variant (rs356182) was among the top PAR signals for European, African/Admixed, and Latino genetic ancestries though more strongly associated with disease in Europeans (Table S7). Moreover, the rs10513789 variant at the *MCCC1* locus was found among the top signals in the African/African Admixed, European, and East Asian genetic ancestry populations; and *VPS13C* rs2251086 ranked among the highest PAR estimates in all groups (Figure 2)(Table S4).

## Discussion

This study proposed the application of PAR with GWAS summary statistics to prioritize both homogeneous and heterogeneous genetic targets across genetic ancestry strata. While PAR fractions for modifiable risk factors in neurodegenerative diseases have been previously assessed^13^, to our knowledge, this is the first study to apply it in a context for AD/PD-related genetic variation.

In AD, East Asian and European ancestry populations exhibited the highest *APOE* PAR estimates, underscoring the well-established risk associated with *APOE4* in these groups^5,6^. *TRANK1*, a novel locus identified in a GWAS on African Americans and deemed crucial in populations of African descent^9^, ranked among the top PAR loci in the African American genetic ancestry population. Other notable loci included *TSPAN14*, for Europeans and African American genetic ancestry individuals, and *PICALM* in all groups, both of which have been suggested as potential therapeutic targets due to their role in microglia activation^14^.

In PD, *SNCA, MCCC1*, and *VPS13C*, and *MAPT* had the highest PAR across multiple strata. Three of these loci are multi-signal with *MAPT* having strong causal variants in Europeans. The *GBF1* locus tagged by the rs10748818 variant had the highest PAR estimate for the African/African Admixed genetic ancestry population. The *HLA-DRB5* locus, which has been previously suggested as a potential genetic overlap between inflammatory bowel disease and PD^15^, exhibited high PAR for the rs112485576 variant. PAR estimates for *GBA1* coding variants were low for most genetic ancestry strata except for the population-specific intronic variant rs3115534, showing the strongest disease association for the African/African Admixed genetic ancestry samples. Similarly, lower PAR estimates for the *LRRK2* rs76904798 variant tagging p.G2019S across ancestries are expected since this variant is relatively less frequent compared to other PD GWAS loci resulting in a smaller population attributable influence^8^. These results highlight key variable risk factors signals and suggest targeted therapies in these strata, underscoring the need to tailor treatments to distinct genetic profiles to potentially increase efficiency in future studies.

This study aimed to estimate PAR for genetic loci associated with AD and PD risk across different ancestries. We note that there are important limitations with the current report, including that low sample sizes, varying LD patterns, and admixture in global datasets, especially in non-European ancestries, may lead to underpowered analyses, hindering generalizability with PAR. Obtaining granular subpopulation summary statistics, including whole genome sequencing data in future GWAS, and expanding on genetic and environmental risk factors could enhance future applications of PAR, making for more efficient discovery analyses by leveraging natural allele frequency variability. Overall, our results demonstrate that the PAR for two common and representative neurodegenerative diseases vary across populations, which may have implications for global public health.

## Data Availability

Data (DOI 10.5281/zenodo.13755496, release 8) used in the preparation of this article were obtained from Global Parkinson's Genetics Program (GP2). GP2 is funded by the Aligning Science Across Parkinson's (ASAP) initiative and implemented by The Michael J. Fox Foundation for Parkinson's Research (https://gp2.org). For a complete list of GP2 members see https://gp2.org.
Information about data access and all scripts for analyses are publicly available on GitHub (DOI 10.5281/zenodo.13774455; https://github.com/GP2code/PAR-ADPD/).

## Acknowledgments

We thank all the participants who contributed to this study. This work was supported in part by the Intramural Research Program of the NIH, the National Institute on Aging (NIA), the NIH, the US Department of Health and Human Services (project number ZIAAG000534), the National Institute of Neurological Disorders and Stroke (NINDS), and NHGRI. We thank Paige Brown Jarreau for her meticulous editing of this manuscript.

Data (DOI 10.5281/zenodo.13755496, release 8) used in the preparation of this article were obtained from Global Parkinson’s Genetics Program (GP2). GP2 is funded by the Aligning Science Across Parkinson’s (ASAP) initiative and implemented by The Michael J. Fox Foundation for Parkinson’s Research (https://gp2.org). For a complete list of GP2 members see https://gp2.org.

## Funding

This research was supported in part by the Intramural Research Program of the NIH, National Institute on Aging (NIA), National Institutes of Health, Department of Health and Human Services; project number ZIAAG000534, as well as the National Institute of Neurological Disorders and Stroke (IFM received R01NS132437-01A1). C.C.C is funded by a Canadian Institutes of Health Research Vanier Scholarship.

## Author contributions

Data was made available in part through the Global Parkinson’s Genetics Program (GP2). L.J. ran analyses for the project. L.J., C.C.C., A.F.S., and S.B.C. contributed equally to interpretation of results and manuscript writing. M.B.M., H.I., M.A.N., A.J.N., C.B., A.S., I.M., M.R.C., and S.B.C. facilitated the review process. The project was supervised by H.I., M.A.N., and senior author S.B.C.

## Potential conflicts of interest

L.J., H.I, M.B.M, and M.A.N.’s participation in this project was part of a competitive contract awarded to DataTecnica LLC by the National Institutes of Health to support open science research. M.A.N. also owns stock in Clover Therapeutics and Neuron23 Inc.

## Notes

### Summary of Updates

Methods revised to include additional analyses; Title, along with the rest of the manuscript, contains minor re-wording; Figure 1 and Figure 2 include re-worded headers; Supplemental files updated; Author list updated.

## References

1. Ji, Z., Chen, Q., Yang, J. et al. Global, regional, and national health inequalities of Alzheimer’s disease and Parkinson’s disease in 204 countries, 1990–2019. Int J Equity Health 23, 125 (2024). 10.1186/s12939-024-02212-5

2. Foo JN, Tan LC, Irwan ID, et al. Genome-wide association study of Parkinson’s disease in East Asians. Hum. Mol. Genet. 2017;26(1):226–232.

3. Rizig M, Bandres-Ciga S, Makarious MB, et al. Identification of genetic risk loci and causal insights associated with Parkinson’s disease in African and African admixed populations: a genome-wide association study. Lancet Neurol. 2023;22(11):1015–1025.

4. Loesch DP, Horimoto ARVR, Heilbron K, et al. Characterizing the Genetic Architecture of Parkinson’s Disease in Latinos [Internet]. Ann. Neurol. 2021;Available from: http://dx.doi.org/10.1002/ana.26153

5. Shigemizu D, Mitsumori R, Akiyama S, et al. Ethnic and trans-ethnic genome-wide association studies identify new loci influencing Japanese Alzheimer’s disease risk. Transl. Psychiatry 2021;11(1):151.

6. Bellenguez C, Küçükali F, Jansen IE, et al. New insights into the genetic etiology of Alzheimer’s disease and related dementias. Nat. Genet. 2022;54(4):412–436.

7. Lake J, Warly Solsberg C, Kim JJ, et al. Multi-ancestry meta-analysis and fine-mapping in Alzheimer’s disease. Mol. Psychiatry 2023;28(7):3121–3132.

8. Nalls MA, Blauwendraat C, Vallerga CL, et al. Identification of novel risk loci, causal insights, and heritable risk for Parkinson’s disease: a meta-analysis of genome-wide association studies. Lancet Neurol. 2019;18(12):1091–1102.

9. Kunkle BW, Schmidt M, Klein H-U, et al. Novel Alzheimer Disease Risk Loci and Pathways in African American Individuals Using the African Genome Resources Panel: A Meta-analysis. JAMA Neurol. 2021;78(1):102–113.

10. Jonson C, Levine KS, Lake J, et al. Assessing the lack of diversity in genetics research across neurodegenerative diseases: a systematic review of the GWAS Catalog and literature [Internet]. medRxiv 2024;Available from: http://dx.doi.org/10.1101/2024.01.08.24301007

11. Wang T, Hosgood HD, Lan Q, Xue X. The Relationship Between Population Attributable Fraction and Heritability in Genetic Studies. Front. Genet. 2018;9:352.

12. International Parkinson Disease Genomics Consortium, Nalls MA, Plagnol V, et al. Imputation of sequence variants for identification of genetic risks for Parkinson’s disease: a meta-analysis of genome-wide association studies. Lancet 2011;377(9766):641–649.

13. Bothongo PLK, Jitlal M, Parry E, et al. Dementia risk in a diverse population: A single-region nested case-control study in the East End of London. Lancet Reg Health Eur 2022;15:100321.

14. Yang X, Wen J, Yang H, et al. Functional characterization of Alzheimer’s disease genetic variants in microglia. Nat. Genet. 2023;55(10):1735–1744.

15. Kars, M.E., Wu, Y., Stenson, P.D. et al. The landscape of rare genetic variation associated with inflammatory bowel disease and Parkinson’s disease comorbidity. Genome Med 16, 66 (2024). 10.1186/s13073-024-01335-2

